# A generalized ODE susceptible-infectious-susceptible compartmental model with potentially periodic behavior

**DOI:** 10.1101/2022.06.10.22276255

**Authors:** Scott Greenhalgh, Anna Dumas

**Affiliations:** Department of Mathematics, Siena College, 515 Loudon Road, Loudonville, NY, 12211

**Keywords:** infectious period, duration of infection, gonorrhea, integral equations, differential equations

## Abstract

Differential equation compartmental models are crucial tools for forecasting and analyzing disease trajectories. Among these models, those dealing with only susceptible and infectious individuals are particularly useful as they offer closed-form expressions for solutions, such as the logistic equation. However, the logistic equation has limited ability to describe disease trajectories since its solutions must converge monotonically to either the disease-free or endemic equilibrium, depending on the parameters. Unfortunately, many diseases exhibit periodic cycles, and thus, do not converge to equilibria. To address this limitation, we developed a generalized susceptible-infectious-susceptible compartmental model capable of accurately incorporating the duration of infection distribution and describing both periodic and non-periodic disease trajectories. We characterized how our model’s parameters influence its behavior and applied the model to predict gonorrhea incidence in the US, using Akaike Information Criteria to inform on its merit relative to the classical SIS model and an SIS model with a time-varying recovery rate. The significance of our work lies in providing a novel susceptible-infected-susceptible model whose solutions can have closed-form expressions that may be periodic or non-periodic depending on the parameterization. Our work thus provides disease modelers with a straightforward way to investigate the potential periodic behavior of many diseases and thereby may aid ongoing efforts to prevent recurrent outbreaks.

## 1. Introduction

A rare few ODE compartmental models have solutions that are closed-form expressions. Most view this list as the class of susceptible-infectious and susceptible-infectious-susceptible (SIS) models, which amount to some form of the logistic growth equation [1,2]. Less well-known are the infectious-recovered (IR) models [3], which are akin to exponential growth, and thereby also deserve inclusion. Historically, these simple models have proven to be of great utility in the study of disease dynamics. For instance, SIS models are commonly used as a first means to predict the total number of people that an epidemic will infect, through the use of the associated final size equation [2,4]. They are also used in the framework to estimate the doubling-time of an epidemic [5] and can be applied to determine the vaccination level required to cause disease burnout [6]. While IR models may have fewer applications, they are infamously quoted in media and literature [7], as “exponential growth” is synonymous with most pandemics. Consequently, lay audiences and novice disease modelers are far more likely to be familiar with the behavior of IR models, at least in comparison to other model types.

Inarguably, SI, SIS, and IR differential equation models and their applications have inspired many current and future disease modelers. These models have limitations though. For instance, their ability to predict the trajectory of diseases is limited, as all trajectories monotonically converge to an equilibrium. Thus, these models cannot predict any form of recurrent epidemics, such as seasonal influenza, measles, and many sexually transmitted infections, among others. While current extensions of these models can address this issue, they do so at the cost of losing the closed-form expressions that make these elementary models convenient.

Here, we propose a new take on the SIS model. We derive our model starting from the general formulation of SIS models as a system of integral equations [8,9] under a general assumption placed on the duration of infection distribution. For particular cases of this class of distributions, we show that our generalized susceptible-infected-susceptible model (gSIS) reduces to the traditional SIS model [10–12] with constant coefficients, and the IR model, respectively. In addition to these subclasses, we also show that our SIS model truly reflects the removal of person-days of infection, and is nonautonomous directly because of the assumptions imposed on the duration of infection. Furthermore, while many other works may have tackled SIS models formulated as logistic growth equations [13–16], we illustrate how to connect the time-varying rates directly to the duration of infection, and transmission rate. To achieve this, we propose that the duration of infection distribution belongs to a particular family of generalized exponential distribution [17], rather than the classical exponential distribution that is commonly inappropriate for describing the duration of infection [18–22].

As proof of concept, we apply our generalized susceptible-infectious-susceptible (gSIS) model to predict gonorrhea incidence in the US. Gonorrhea is a nationally reportable sexually transmitted disease [23,24], which is typically caused by the spread of bacteria during sexual contact [25], although mother-to-child transmission is possible. While medical treatment is available, gonorrhea infection confers little to no immunity [26], with re-infection common after subsequent exposures [27]. However, despite this dynamic of infection, return to susceptibility, and re-infection, trends in gonorrhea infection can appear oscillatory [28], rather than convergent to a disease-free or endemic equilibrium, which casts doubt on the applicability of traditional SIS models for predicting disease trends. So, we apply our gSIS model to capture the oscillatory trajectory of gonorrhea in the US, all the while maintaining the convenient properties expected of traditional SIS models.

## 2. Methods

In what follows, we illustrate a gSIS model, as formulated by a system of ODEs. We derive the model starting from a system of Volterra integral equations, imposing the assumption that the infectious period belongs to a class of generalized exponential distributions. We then proceed to reduce the model to a logistic growth equation with time-varying parameters, illustrate the closed-form solutions for all special parameter values (Table 1), and completely characterize the stability properties of the system, including the potential occurrence of periodic cycles.

**Table 1.**
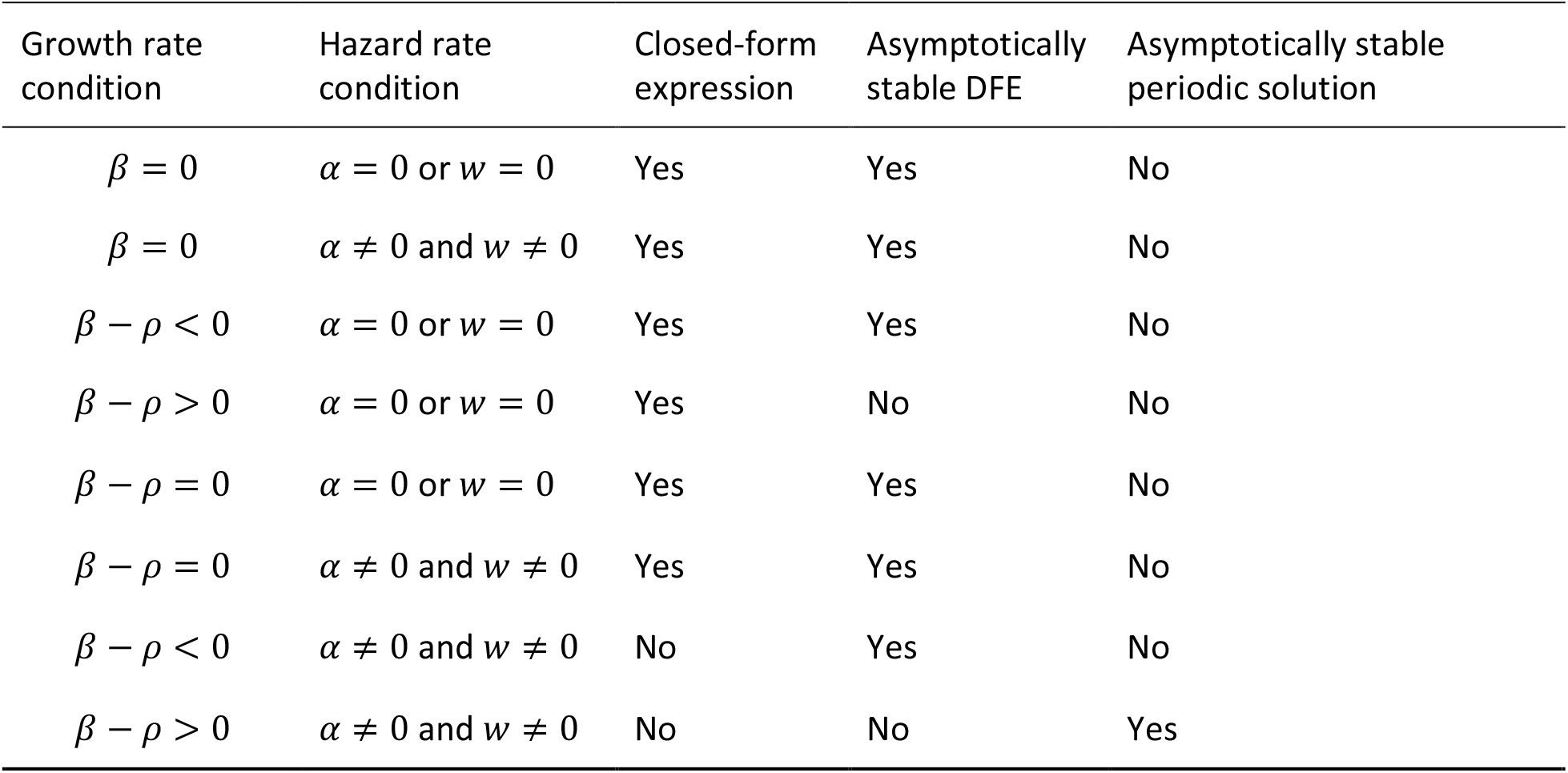
Summary of gSIS model stability conditions and properties.

### 2.1 Generalized differential equation compartmental models

Classically, one of the most general forms of compartmental models is those formulated as a system of Volterra integral equations [8,10–12,29,30]. For this classical compartmental model, the proportion of susceptible individuals is denoted as *s*(*t*), and the proportion of infected individuals is denoted as *i*(*t*), [8,30], where *i*_0_(*t*) characterizes the proportion of infected individuals at the beginning of the epidemic. The relation between compartments is given by

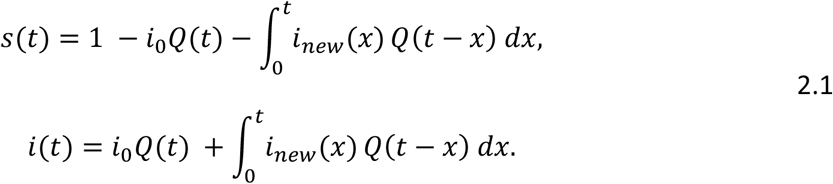

Here *i*_*new*_(*x*) is the rate of new incidence at time *x, i*_0_ is the initial proportion of infected individuals, and *Q*(*t* − *x*) is the survival function associated to the infectious period distribution, which describes the proportion of individuals that remains infectious at time *t* given infection occurred at time *x*.

Classically, the formulation of *i*_*new*_(*x*) is due to a simplification of the law of mass action [31–33], namely

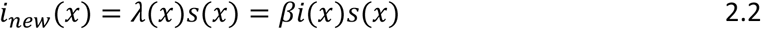

whereby new infections are proportional to the product of the proportions of infected and susceptible individuals.

From 2.1, one can obtain the traditional differential equation SIS model (with recovery rate *γ*) by imposing *i*_*new*_(*x*) = *βi*(*x*)*s*(*x*) and *P*(*t* − *x*) = *e* ^−*γ*(*t*−*x*)^, differentiating 2.1 with respect to *t*, and then substituting the remaining integral terms with the appropriate multiple of equations from 2.1 respectively. Alternatively, by imposing 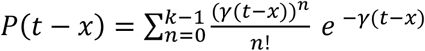, i.e. an Erlang distributed duration of infection, one obtains the SI^k^S model [22], where the state of infection has been subdivided into *k* stages.

Given *i*(*x*) represents the proportion of infected individuals from different initial moments of infection a seldom acknowledged issue is whether the infectious period distribution is still appropriate to describe survival. Replacing this distribution with the related duration of infection distribution would likely be more appropriate, which implies that

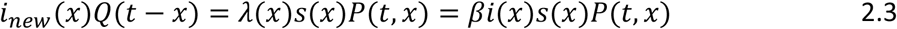

where by definition *P*(*t*, 0) = *Q*(*t*).

We thus arrive at more flexible analog of system 2.1, namely

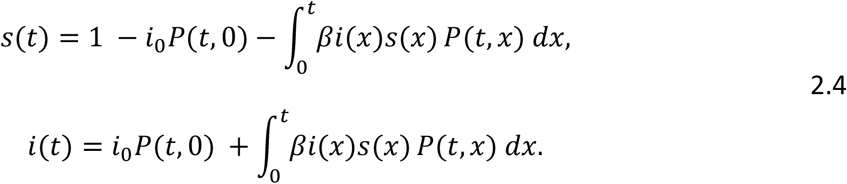

While many forms of *P*(*t, x*) are appropriate, we assume it follows a non-homogeneous analog of the exponential distribution, specifically

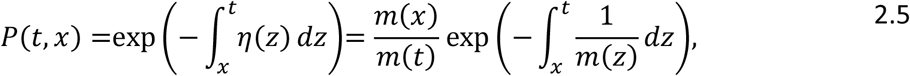

where *η*(*t*) = (*m*′(*t*) + 1)/*m*(*t*) is the hazard rate and *m*(*t*) is the mean residual waiting-time of the duration of infection. It also follows that the equilibrium distribution (the limiting distribution of forward recurrence times) [34] is defined as

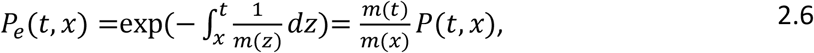

which allows system 2.4, upon multiplication by *m*(*t*), to be rewritten as

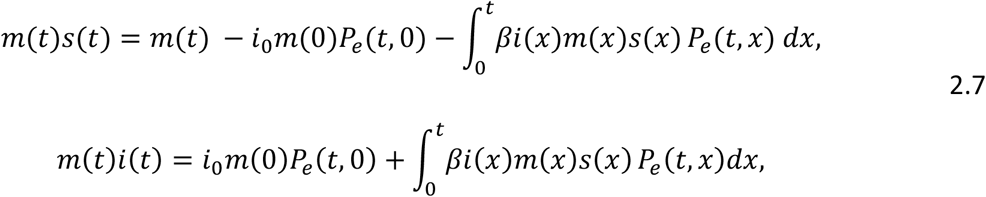

If we further multiply by the total population, *N*, equations 2.7 describe person-days susceptible to infection and infected, where the survival of individuals infected at time *x* is based on the equilibrium distribution *P*_*e*_(*t, x*).

Differentiating 2.7, substituting their multiple by 1/*m*(*t*) and then isolating for *s*′(*t*) and *i*′(*t*), respectively, we arrive at the time-varying SIS model

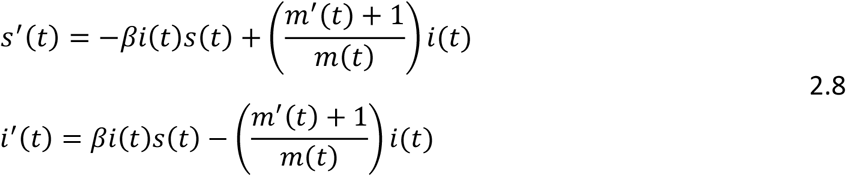

An alternative to a compartmental model based on incidence or proportion of infected individuals is to instead consider person-days of infection (the number of infectious people multiplied by their average duration of infection) from the onset. For this compartmental model, the number of person-days of infection is denoted as *D*(*t*), where *D*_*new*_(*t*) characterizes the person-days of infection from newly infected individuals who are initially infectious at time *t*. Thus, the number of person-days of infection can be modelled as

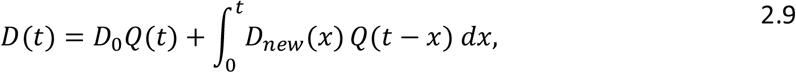

where *D*_0_ is initial person-days of infection from infectious individuals at the start of the epidemic, and once again *Q*(*t* − *x*) is the survival function associated to the infectious period distribution. Denoting *N* as the total population, by an assumption similar to 2.3, we have that

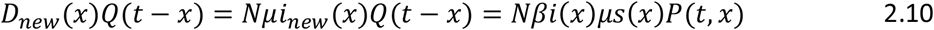

or

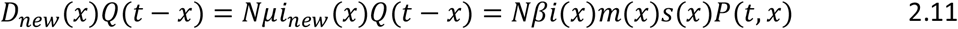

The distinction between 2.10 and 2.11 is in the choice of *μ* (which equals *m*(0)) versus *m*(*x*). Biologically, *m*(*x*)*i*(*x*)*P*(*t, x*) describes the survival of the exact number of person-days of infection at time *x*, whereas *μi*(*x*)*P*(*t, x*) is likely an over or under estimate, as it does not account for the variation in person-days of infection due to the different initial times of infection of infected individuals.

Further recognizing that person-days of infection is the product of the number of infectious individuals and their average duration of infection, it follows that

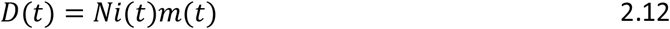

Assuming 2.11 and 2.12, it follows that equation 2.9 becomes

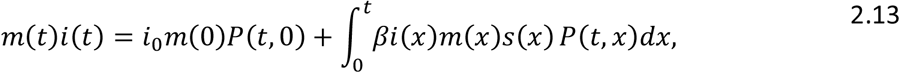

where we now include

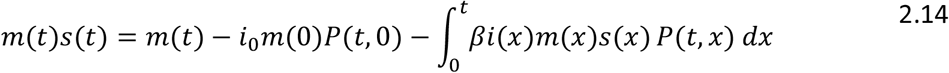

to ensure the conservation of population.

The structure of equations 2.13 and 2.14 is similar to that of system 2.7, the difference amounting to the choice of *P*(*t, x*) rather than *P*_*e*_(*t, x*). The motivation to choose *P*(*t, x*) over *P*_*e*_(*t, x*) stems from their definitions, as *P*(*t, x*) exactly describes the removal of the proportion of infection *i*(*t*) with time-varying average duration of infection *m*(*t*), whereas *P*_*e*_(*t, x*) does so only in the equilibrium sense.

Multiplying 2.13 through by *P*(*τ, t*) and using the property that *P*(*τ, t*)*P*(*t, x*) = *P*(*τ, x*), we can predict the survival of person-days of infection from any moment *t* (ignoring any future transmission) as

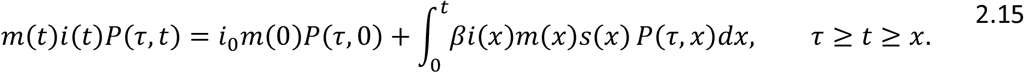

To obtain a generalized differential equation compartmental model (GDECM), we differentiate equation 2.15 with respect to *t* and divide through by *P*(*τ, t*) to obtain (*i*(*t*)*m*(*t*))_′_, and then apply the conservation of population to obtain (*s*(*t*)*m*(*t*))_′_ :

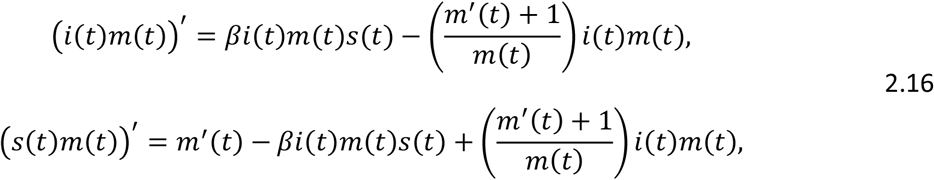

or if we isolate for *s*′(*t*) and *i*′(*t*) specifically, we arrive at

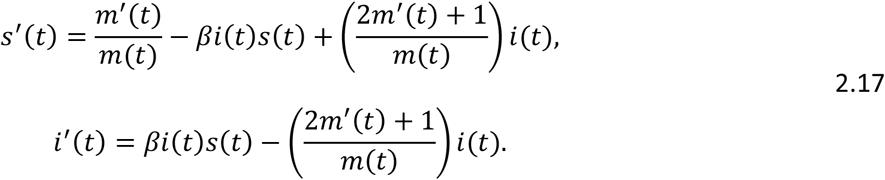

The difference between equations 2.17 and 2.8 ultimately amounts to the coefficient of *m*′(*t*) in *i*′(*t*). Changing this coefficient from 1 in 2.8 to 2 in 2.17 ensures that the removal of person-days of infection reflects their true survival, rather than an equilibrium approximation.

### 2.2 The periodic hazard rate and mean residual waiting-time

The hazard rate and mean residual waiting-time functions are constructs often used in survival and reliability analysis to describe the frequency of events and the time remaining in a given state, respectively, after an elapsed period [35]. The hazard rate function also characterizes [35,36] the mean residual waiting-time through

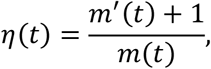

and is often either non-increasing or non-decreasing [37]. Recent work, however, illustrates the utility of more flexible functional forms. Specifically, hazard rates with a bathtub [37], upside-down bathtub [38], or roller-coaster [39] shape have been applied broadly in various contexts [40]. Here, we consider potentially periodic hazard rates, depending on model parameterization [17].

Perhaps the simplest probability density function (PDF) with a periodic hazard rate is the exponential distribution augmented with a time-dependent cosine rate parameter:

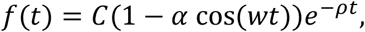

where *α* ∈ (−1,1), *w* ≥ 0, *ρ* > 0, *t* ≥ 0, and 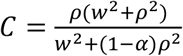.

Given the PDF the cumulative distribution function (CDF) [17,41] is

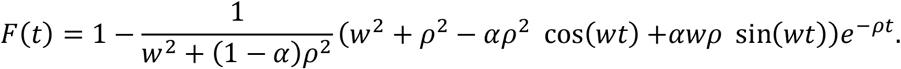

It follows that the hazard rate is

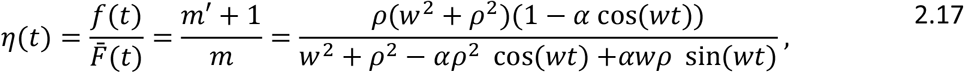

Where 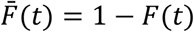 is the survival function.

Furthermore, through solving the relation *η*(*t*) = (*m*′(*t*) + 1)/*m*(*t*) with 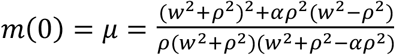, we also have that the mean residual waiting-time is

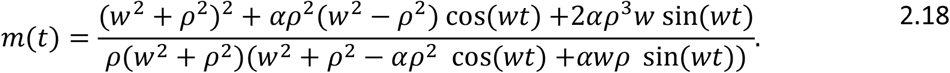

The average recovery rate at time *t* naturally follows as

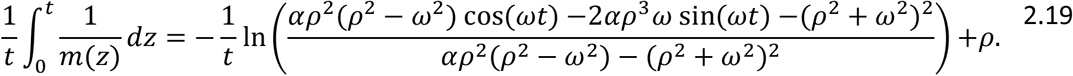

### 2.3 GDECM solutions and properties

Using *s*(*t*) + *i*(*t*) = 1 and the assumption that *m*(*t*) is time-dependent, we have that system 2.16 reduces to a Bernoulli equation [42] of the form

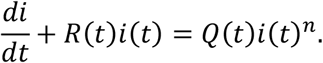

Thus, the nonlinear ODEs can be transformed through *i*(*t*) = 1/*y*(*t*) into the first-order linear non-constant coefficient ODEs of the form

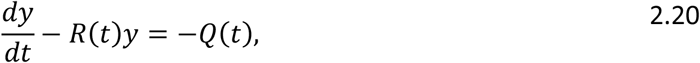

As the transformed ODE is first-order and linear it can be solved by integrating factors.

For system 2.16 it follows that 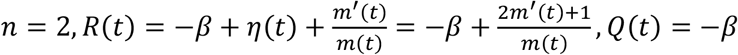, thus the integrating factor is

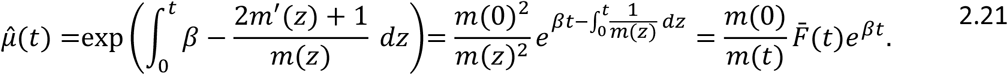

Solving yields the ODE

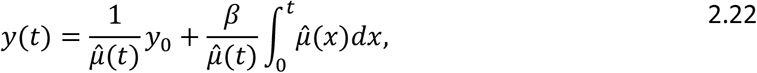

or upon reversing the transformation,

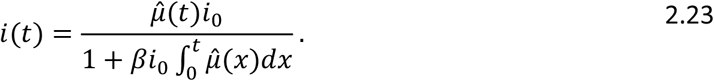

We now proceed to illustrate that 2.16 includes SIS, and IR classes of compartmental models, and completely classify its stability behavior.

#### 2.3.1 The case when *β* = 0, and *w* = 0 or *α* = 0

In this case, equation 2.16 reduces to an IR model, as described by an exponential decay. The solution in general is given by

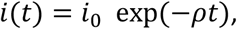

which decays at the rate *ρ* to the disease-free equilibrium (DFE), *i*(*t*) = *i*_*DFE*_ = 0.

#### 2.3.2 The case when *β* = 0, and *w* ≠ 0 or *α* ≠ 0

Similar to section 2.3.1, this model also reduces to an IR model, but the decay rate is time-dependent. The general solution for 2.16 is

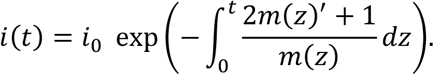

The solution possesses closed-form expressions when 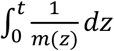 is integrable. It also follows that solutions decay to the DFE, as *m*(*t*) > 0.

#### 2.3.3 The case when *β* − *ρ* ≠ 0, and *w* = 0 or *α* = 0

In this case, the duration of infection distribution reduces to an exponential distribution. When *β* − *ρ* ≠ 0, with *w* = 0 or *α* = 0, it follows that *m*(*t*) = 1/*ρ*. So, equation 2.16 reduces to an SIS model, or equivalently a constant coefficient logistic differential equation. Thus, we have that

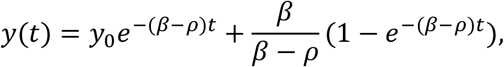

which implies that

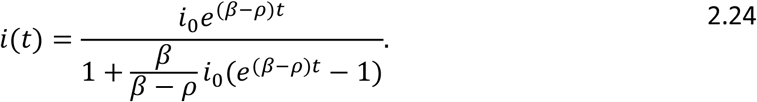

The stability of each equilibrium of 2.16 is determined by the sign of *β* − *ρ*, or equivalently the basic reproductive number, as calculated through the next-generation approach [43], which is given by

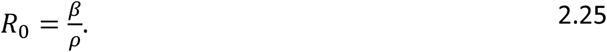

It follows that the DFE is stable when *R*_0_ < 1 and the endemic equilibrium is stable when *R*_0_ > 1. Note, the stability for *R*_0_ = 1 cannot be determined here, as the assumption *β* − *ρ* ≠ 0 implies *R*_0_ ≠ 1.

#### 2.3.4 The case when *β* − *ρ* = 0, and either *w* = 0 or *α* = 0

Under such conditions, *m* = 1/*ρ*, and 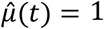. It follows that

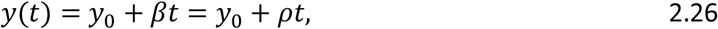

and thus

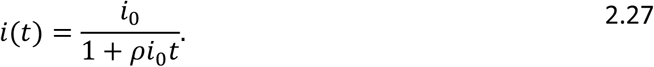

Given that *ρ* > 0, it follows that the DFE is stable, as *i*(*t*) → 0 when *t* → ∞.

#### 2.3.5 The case when *β* − *ρ* = 0, *w* ≠ 0, *α* ≠ 0

When *β* − *ρ* = 0, the solution of the integrating factor is

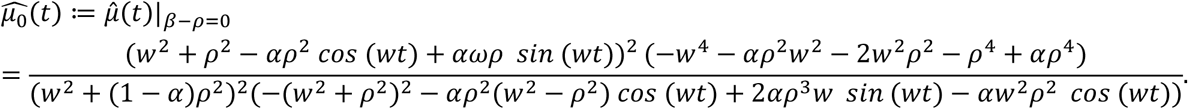

Furthermore, it follows that the integral of 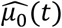 also has a closed-form expression, as for integer *n* > 0 we have that

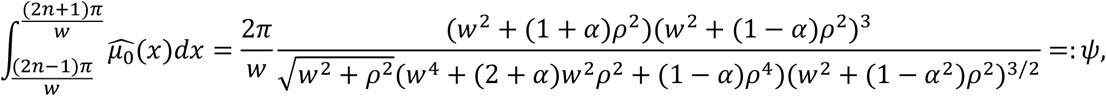

and

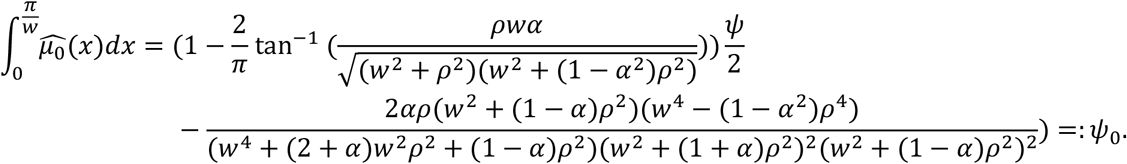

For 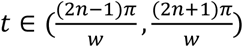 the integral of 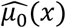 is

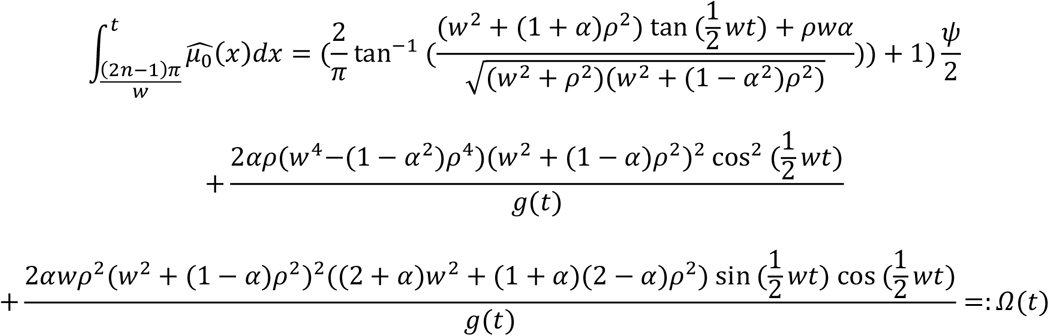

where

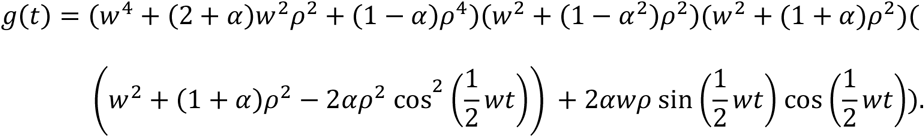

Thus, provided 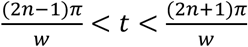, we have that

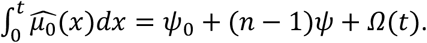

It follows that *i*(*t*) has a solution represented by a closed-form expression because 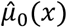 and 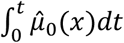 have such a feature. Furthermore, given that 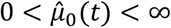 is periodic, we have that 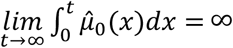, which causes 2.11 to converge to the DFE.

#### 2.3.6 The case when *β* − *ρ* ≠ 0, *w* ≠ 0, *α* ≠ 0

For this case the solutions are up to the quadrature of 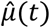. To understand the qualitative behavior of *i*′(*t*), we first consider its formulation as a logistic equation with time-varying parameters:

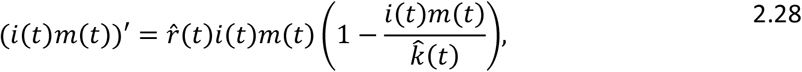

where 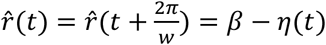 and 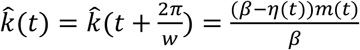.

To demonstrate the stability properties of 2.28, it suffices to provide conditions so that 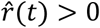 and 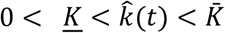. As *β* > 0, *m*(*t*) > 0, and 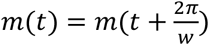, this problem reduces further to investigating 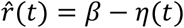.

The critical points of 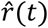 occur when

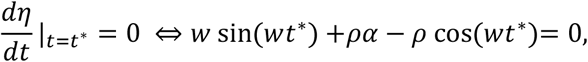

Furthermore, from *t*\*, a local minimum occurs when

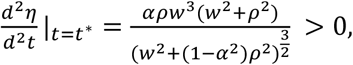

and a local maximum when

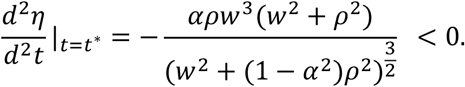

It follows that

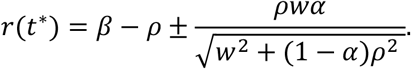

Thus, if

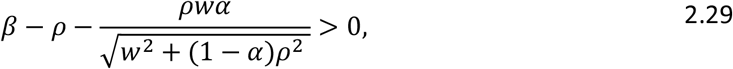

then 2.28 has two non-negative periodic solutions [14,15], with one being the DFE. The second periodic solution arises because there exists 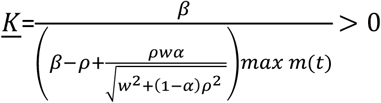, and 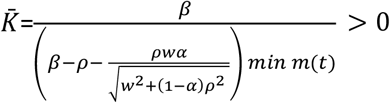. To elaborate, because 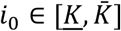 we have that 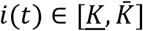, furthermore, if *i*_0_ < *<u>K</u>* then *i*′(*t*) > 0, and when 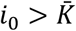 then *i*′(*t*) < 0. Thus, 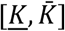 acts as a trapping region for *i*(*t*), and because there does not exist an equilibrium solution in 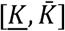, and the dimension of the system is 1, *i*(*t*) must be periodic. It also follows that this second periodic solution satisfies the criteria to be asymptotically stable [15].

To address when 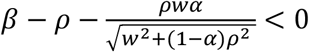, we first assume there exists *ϵ* > 0 so that *β* = *ρ* + *ϵ*. It follows that

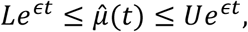

where 0 < *L* ≤ *U* are constants.

Thus, given *i*_0_ > 0, from 2.23 we have that

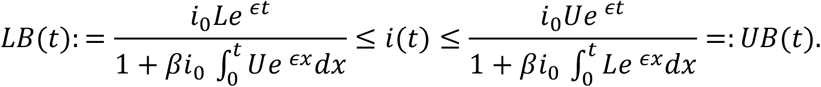

Taking the limit as *t* → ∞ and applying l’Hôpital’s rule, we obtain the bounds

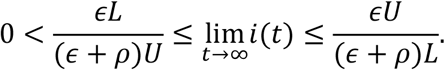

We also have that

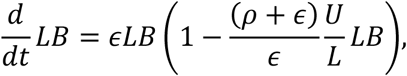

and

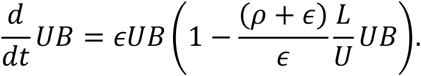

Naturally, this implies that

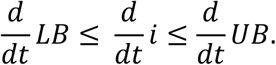

As a consequence of these bounds, if 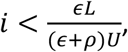, then 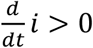, and if 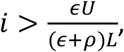, then 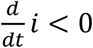. Similar to when *r*(*t*) is strictly positive, 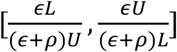 acts as a trapping region. Therefore, provided *β* − *ρ* > 0, there exists a second periodic solution, which is asymptotically stable (Supplementary Information). Next, we consider *ϵ* > 0 so that *β* = *ρ* − *ϵ*. It follows once again that constants exist so that

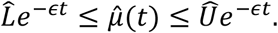

Given these bounds on the integrating factor 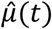, we have that

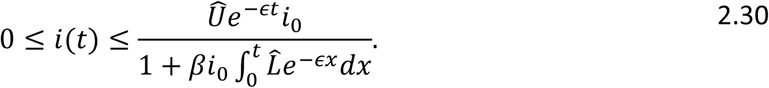

Taking the limit, we have that 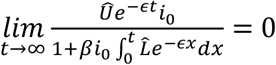, and thus *i*(*t*) converges to the DFE.

## 3. Application of the methodology to gonorrhea in the United States

To illustrate the utility of our gSIS model, we apply it, the traditional SIS model, and an SIS model with a time-varying recovery rate to predict gonorrhea incidence in the United States. We estimate parameters by minimizing the least-square error of model predictions with historical data on gonorrhea incidence [23,24], as well as the literature [44,45]. Specifically, we estimate the average duration of infection based on an average of 1.5 days from gonorrhea exposure to infectiousness [44], an incubation period of approximately 7.5 days [23], and an average of 7.4 days from symptom onset to visiting a medical professional for treatment. Altogether, this implies the average duration of infection is

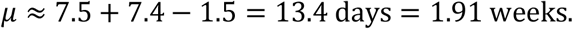

Using this estimate, we reduce the number of parameters to estimate by imposing that *ρ* satisfies

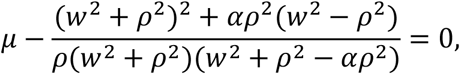

where *w* and *α* are determined by minimizing the least-square error between model predictions and gonorrhea incidence data for the gSIS and time-varying SIS model or set to zero for the traditional SIS model. Details of additional parameters, including the transmission rate, are available in Table 2.

**Table 2.** Parameters, base values, and sources.

| Parameter | Constant | Value |
| --- | --- | --- |
| Population size (10000s) | $N$ | |
| Traditional SIS |  | 48.4 |
| Time-varying SIS |  | 53.0 |
| gSIS |  | 42.4 |
| Duration of infection (in absence of periodicity) | $1/\rho$ | |
| Traditional SIS |  | 1.91 weeks |
| Time-varying SIS |  | 1.98 weeks |
| gSIS |  | 1.97 weeks |
| Transmission rate | $\beta$ | |
| Traditional SIS |  | 0.541/week |
| Time-varying SIS |  | 0.519/week |
| gSIS |  | 0.523/week |
| Period of the epidemic | $2\pi/w$ | |
| Traditional SIS |  | Undefined |
| Time-varying SIS |  | 41.4 weeks |
| gSIS |  | 39.6 weeks |
| Magnitude of seasonality | $\alpha$ | |
| Traditional SIS |  | 0 |
| Time-varying SIS |  | -0.275 |
| gSIS |  | -0.241 |

Given the estimate of model parameters, the (time-varying) average duration of infection varies between 1. 84 weeks and 2.12 weeks for the gSIS model, between 1.83 and 2.14 weeks for the time-varying SIS model, and remains constant at 1.91 weeks for the SIS model (Figure 1). For the models, we have that the DFE is unstable, as 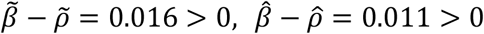, and 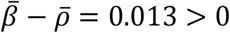 for the gSIS, SIS, and time-varying SIS models, respectively. Using the estimated parameters (Table 2), the gSIS, traditional SIS, and time-varying SIS models predict a peak and trough of 2415-4709, and 2415-4683 new incidences of gonorrhea per week, respectively, over the next 300 weeks (Figure 2). Furthermore, the period of the epidemic for both models was estimated as 39.6 and 41.4 weeks, with an amplitude of seasonality of -0.241 and -0.275, respectively.

**Fig. 1.**
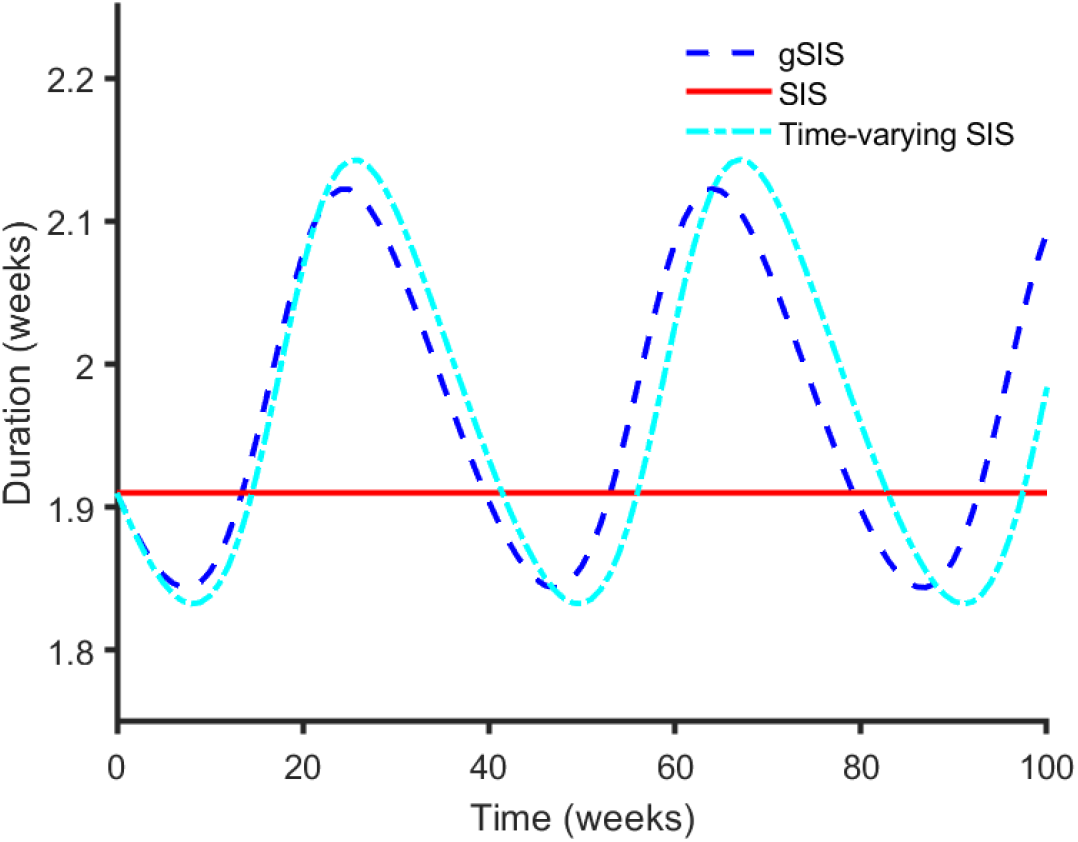
The time-varying average duration of infection. Estimate of average duration of infection of gonorrhea at time *t* for the gSIS model with 1/*ρ* = 1.97 weeks, 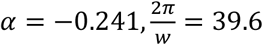 weeks (blue dashed curve), the SIS model with 1/*ρ* = 1.91 weeks, *α* = *w* = 0 (red solid curve), and the time-varying SIS model with 1/*ρ* = 1.98 weeks, 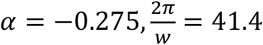 (cyan dash-dot curve).

**Fig. 2.**
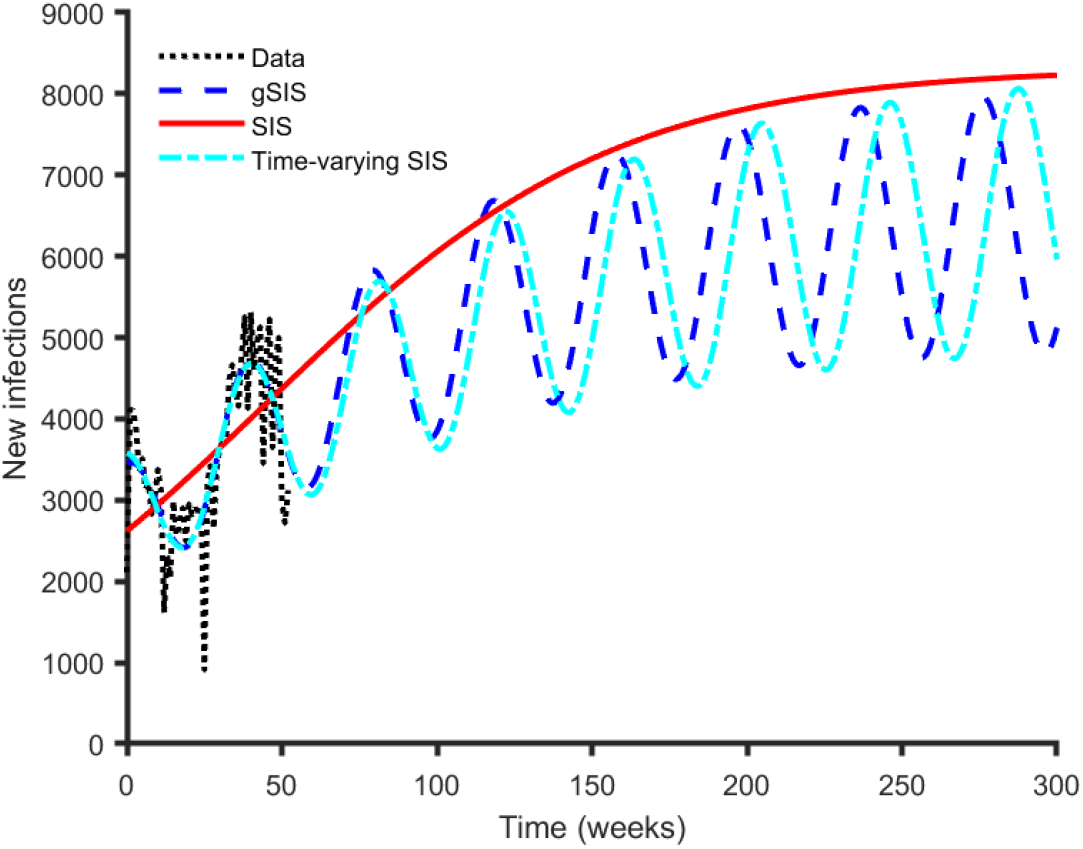
Model fit and predictions of new gonorrhea infections. The trajectory of new gonorrhea infections based on data (black dotted curve), the gSIS model with 1/*ρ* = 1.97 weeks, 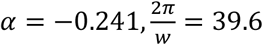 weeks (blue dashed curve), the SIS model with 1/*ρ* = 1.91 weeks, *α* = *w* = 0 (red solid curve), and the time-varying SIS model with 1/*ρ* = 1.98 weeks, 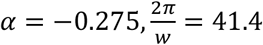 (cyan dash-dot curve).

To inform on the merit of our predictions, we calculated the Akaike information criteria (AIC) [46]. Briefly, AIC is a mathematical method for comparing how well models fit data, relative to one another, which takes into account model complexity [47]. Assuming model solutions are represented by *i*(*t*; *β, ρ, α, w, i*_0_), we define the new incidence as

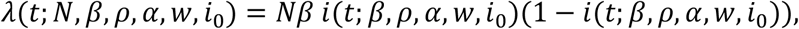

where a subscript of SIS, gSIS, or tvSIS is used to distinguish between cases.

It follows that *AIC*_*gSIS*_ = 656.3, *AIC*_*SIS*_ = 835.5, and *AIC*_*tvSIS*_ = 667.6 (Supplementary Information). Thus, as *AIC*_*gSIS*_ < *AIC*_*tv*_ < *AIC*_*SIS*_, the criterion suggests that gSIS is a more appropriate model for describing the trajectory of gonorrhea.

## 4. Discussion

We demonstrated a novel take on SIS models motivated by their extension to compartmental models that track person-days of infection. For our model, we completely characterized its stability behavior, illustrated its equivalence to two formulations of time-varying logistic growth equations, and demonstrated that all known families of compartmental models that feature solutions with closed-form expressions are special cases. Importantly, this work is also the first to build an SIS compartmental model with time-varying parameters directly from assumptions placed on the duration of infection distribution, while also extending SIS models to potentially feature periodic behavior.

We illustrated a novel SIS model with new solutions that were closed-form expressions beyond the well-known SIS, SI, and IR models. While it remains an open question whether more complex models have such convenient properties, the added flexibility of GDECM should provide a means to obtain additional gSIS models with such features. Furthermore, the GDECM framework may even enable the development of the first SIR models with solutions that have a closed-form expression. If such an SIR model exists, it would imply the existence of an additional conservation law beyond the conservation of population, and therefore could, at least in theory, be tested empirically.

An interesting feature of gSIS and time-varying SIS models is that the existence of periodic solutions requires *β* − *ρ* > 0, or equivalently 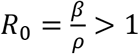. Typically, for autonomous compartmental models, the existence of periodic solutions (when only one endemic equilibrium exists) requires a specific set of parameter values so that *R*_0_ ≈ 1, at least according to Hopf-bifurcation theory [48]. As such, the condition required for periodic solutions of gSIS models suggests oscillatory dynamics are far more likely, which is corroborated by observed trends of many disease trajectories that feature reproductive numbers greater than one.

While our work focuses on gSIS models, it provides an important extension to logistic growth equations with time-varying parameters. To date, the majority of analyses of such logistic models require that the time-varying growth parameter is non-negative for periodic solutions to exist. Here, we showed that it is possible to weaken this condition to periodic rates that are positive more often than negative. Biologically speaking, such a condition seems plausible, at least for diseases or species that reproduce in low numbers frequently, subject to some form of seasonal fluctuations.

The estimated epidemic period of 39.6-41.4 weeks provided by the gSIS and time-varying SIS models closely mirrors the average duration of a school year of approximately 40 weeks (including weekends and holidays). While this similarity could be a coincidence, the high prevalence of gonorrhea among 15-24 year-olds [23,24], and the known role of the school year in the transmission of other diseases [49,50], suggests that the length of the school year may play a prominent role in the periodic behavior of gonorrhea.

A further avenue of research is to consider a gSIS multistrain model that features a mixture of distributions with periodic hazard rates. Specifically, with such a model and distribution it may be possible to ascertain fundamental frequencies of transmission and infection directly from the model’s solution. Subsequently, it may also be possible to determine whether there could be any resonance between strains [51] and how such a thing may contribute to pathogen evolution. Another direction for future work is the merger of gSIS with techniques for predicting the doubling-time of epidemics. While recent work tackles the question of doubling-time for epidemic models in the context of a modified Richards model [5] (a generalization of logistic growth), it stands to reason that such analysis could be applied to gSIS models, given their formulations as logistic growth equations with time-dependent parameters. Relatedly, the solutions with closed-form expression of gSIS should also provide a means to estimate a final size function, akin to a final size equation of autonomous SIS models, that could provide details on the number of infected individuals, given only an initial condition and desired time period.

While this work focuses on a modification of differential equation SIS models to include a time-varying average duration of infection, one could also study gSIS models in the context of integral equations. The added flexibility of such a modification to an integral equation description of disease spread may provide a means to tackle open challenges in disease modeling [52], such as understanding the endemic equilibrium and defining its stability [53].

Extending SIS models to the framework of GDECM carries with it many of the limitations of differential equation compartmental models, with a homogeneously mixed population likely being the most prominent drawback. Furthermore, the extension of SIS to gSIS still may not be sufficient to accurately represent the transmission dynamics of gonorrhea, as the inclusion of treatment, asymptomatic infection, and treatment resistance classes, among others, may be required to accurately capture transmission dynamics. Another drawback of gSIS, at least relative to SIS, is that it requires more abundant information on the average duration of infection, namely how it varies in time. While this requirement may not greatly inhibit the theoretical development and application of this new class of models, it may impede its use in more intensive disease modeling analyses.

In summary, we demonstrated a new type of compartmental models based on extending a simple SIS model to the framework of GDECM. In accomplishing this, we showed how this simple extension adds the potential for rich dynamics, solutions that have closed-form expressions, and more accurately captures the removal of disease from a population. Naturally, generalizing compartmental models further will likely only enhance these features and properties, and thereby provide ample avenues for future investigation.

## Data Availability

The datasets generated during and/or analyzed during the current study are available in a GITHUB repository (Greenhalgh and Dumas 2022), see doi 10.5281/zenodo.6595905

https://zenodo.org/record/6595905#.YqN2BqjMJdg

## Acknowledgments

The authors wish to thank Drs. Troy Day and Emelie Kenney for constructive feedback that greatly improved the clarity of the work

## Supplementary Information

In this appendix, we provide further details on the stability of periodic solutions for our generalized susceptible-infected-susceptible model (gSIS). In addition, we also provide details that demonstrate that the mean residual waiting-time, *m*(*t*), is positive and bounded, and outline our calculation of the Akaike Information Criterion (AIC).

### S.1. Stability conditions of periodic solutions

Consider the gSIS model, as represented by the time-varying logistic growth equation,

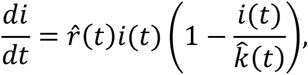

where 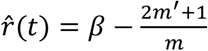, and 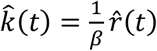, and note that a analogous analysis could also be conducted for the time-varying SIS model by assuming 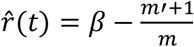, and 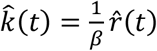.

For the gSIS model, assuming *i*(*t*) is a periodic solution, with period 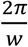, it follows that

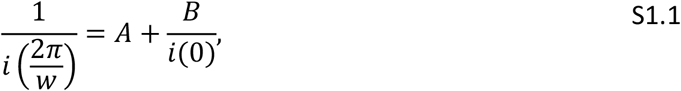

Where

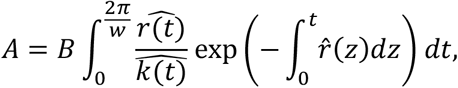

and

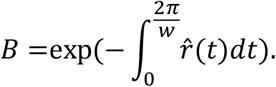

Defining 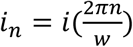, it follows that S1.1 is equivalent to

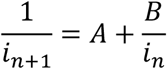

or

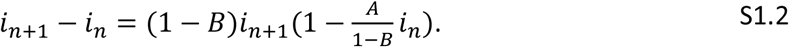

It follows that S1.2 has two equilibria, *i*\* = 0, which corresponds to the DFE of *i*′(*t*), and 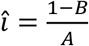, which corresponds to the periodic solution of *i*′(*t*). Linearizing S1.2 about *î*, it follows that î is locally stable provided

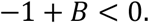

Translating this back to the original system, we have that the periodic solution is locally stable when

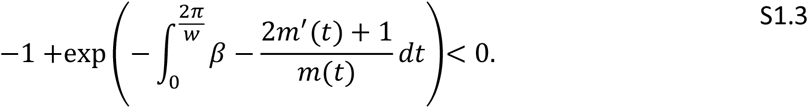

Noting that 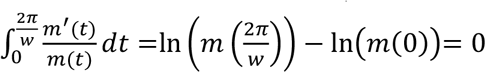, and that 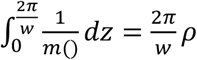 due to the periodicity of *m*(*t*), it follows that S1.3 reduces to

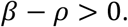

### S.2 Upper and lower bounds on the mean residual waiting-time

Here we show that the mean residual waiting-time, given by

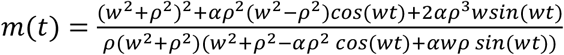

is positive and bounded.

To begin, we determine the critical points of *m*(*t*),

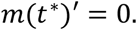

Recalling the hazard rate 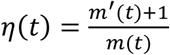, we have that

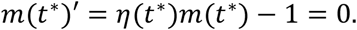

By virtue of *η*(*t*) being positive and bounded [17], it follows that *m* is the same.

### S.3 The calculation of Akaike Information Criteria

Assuming model solutions are represented by *i*(*t*; *β, ρ, α, w, i*_0_), we define new incidence as

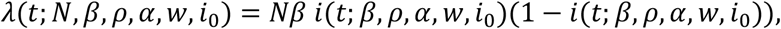

where a subscript of SIS or gSIS is used to distinguish between cases.

It follows for the SIS model that

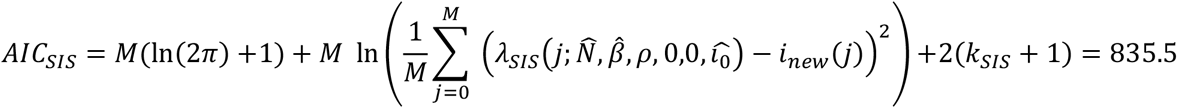

where *M* is the number of data points, *i*_*new*_(*j*) is the observed new incidence on the *j*^*th*^ week, *k*_*SIS*_ = 3 is the number of estimated parameters, and 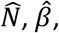 and 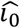 are best estimates of these parameters from the least square minimization procedure.

Similarly, for the time-varying SIS model, it follows that

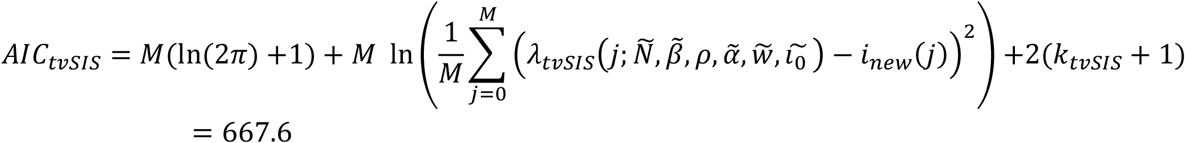

where *k*_*tvSIS*_ = 5 is the number of parameters estimated, and 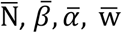 and 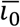 are the best estimates of parameters from the least square minimization procedure for the time-varying SIS model.

Finally, for the gSIS model, we have that

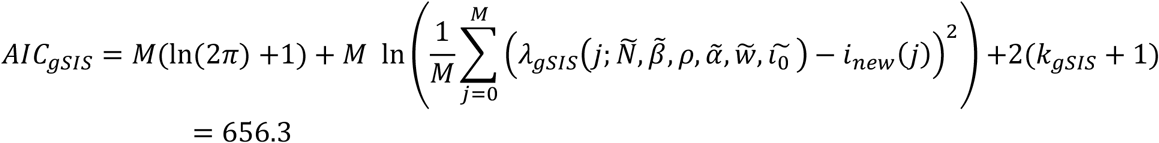

where *k*_*gSIS*_ = 5 is the number of parameters estimated, and 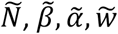 and 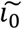 are the best estimates of parameters from the least square minimization procedure for the gSIS model.

